# Rescue Stenting in Mechanical Thrombectomy Refractory Occlusions: A Single Center Multiethnic Cohort

**DOI:** 10.1101/2022.07.30.22278139

**Authors:** Yahia Z Imam, Naveed Akhtar, Saadat Kamran, Pablo Bermejo, Salman Al Jerdi, Ayman Zakaria, Ahmed Own, Satya Patro

**Author notes:** Correspondence: Yahia Z Imam, Neuroscience Institute/Stroke Program, P.O. Box 3050, Doha, Qatar.

## Abstract

**Introduction:** Refractory large vessel occlusion in acute ischemic stroke carries high morbidity and mortality. Rescue stenting is an emerging modality that is increasingly utilized especially in East Asia. We aim to investigate the safety and efficacy of performing rescue stenting in acute stroke patients who had failed mechanical thrombectomy.

**Methods:** This is a retrospective, all-inclusive, observational, descriptive review of the prospectively collected stroke database. Post stenting, an aggressive antiplatelet protocol was followed with glycoprotein IIb/IIIa infusion. Incidence of intracerebral hemorrhage (ICH), recanalization score and favorable prognosis (modified Rankin Score ≤3) at 90 days was used to determine primary outcome. Additionally, comparison was made between patients from the Middle East and North Africa (MENA) region and others.

**Results:** In total, 55 patients were included with 87.3% being male. Mean age was 51.3±11.8 years. This included 32 patients (58.3%) from South Asia, 12 (21.8%) from the MENA region, 9 (16.4%) from Southeast Asia and 2 (3.6%) from other parts of the world. Optimal recanalization (Thrombolysis in cerebral infarction (TICI) scale=2b-3) was achieved in 43 (78.2%); the incidence of symptomatic ICH was 2 (3.6%) and favorable outcome at 90 days was seen in 23 (41.8%). Apart from significantly older age, mean 62.8±13 years (median 69) vs. 48.1±9.3 (mean 49) and coronary artery disease burden 4 (33.3%) vs.1 (2.3%) (p<0.05). Patients from the MENA cohort had similar risk factor profiles, stroke severity, recanalization rates, ICH rates and 90-day outcomes compared to patients from South and Southeast Asia.

**Conclusion:** Rescue stenting showed comparably good outcomes and low risk of clinically significant bleeding in a multiethnic cohort of patients from MENA and South and Southeast Asia.

## Introduction

Mechanical thrombectomy (MT) has revolutionized acute ischemic large vessel occlusion (LVO) stroke treatment(1) with a success rate of 75-80% using the stent-retriever technique. However, some vessels re-occlude after recanalization(1, 2). Reocclusion occurs in 6-24% of patients within the first 24 hours and leads to increased disability and mortality(2, 3). Many factors have been reported to be associated with reocclusion including prior statin use, the occlusion site, presence of residual thrombus or stenosis and the complexity of the MT procedure(3). An important factor associated with reocclusion is the presence of intracranial atherosclerosis(3). Intracranial atherosclerotic disease (ICAD) contributes about 8-10% of strokes in the United States; it is much more prevalent in Asian populations, accounting for 30-50% of ischemic stroke(4, 5).

Although patients with ICAD-related LVO frequently have favorable hemodynamics, collateral scores and smaller cores ; they have poorer response to MT and higher rates of endovascular therapy (EVT) failure or reocclusion (6, 7).This could be related to longer procedure times, presence of hidden stenosis (7) or irritation to the atherosclerotic plaque inducing in-situ thrombosis with multiple passes(6, 7). Therefore, alternative endovascular techniques such as rescue stenting are needed.

Rescue stenting remains controversial. More recent publications (1, 8-14) mainly out of East Asia, have demonstrated the relative safety and efficacy of acute intracranial stenting in patients with refractory LVOs to standard endovascular revascularization techniques such as stent-retriever or direct contact catheter aspiration. Despite being effective, rescue stenting requires treatment with intravenous antiplatelets potentially increasing the risk of intracranial or systemic hemorrhage.

In our cohort of stroke patients, the rate of ICAD etiology is high, likely due to large numbers of patients from South and Southeast Asia in the country(15). Thus, refractory LVOs are not uncommon and rescue acute intracranial stenting has been used for years. The purpose of the current study is to report on the efficacy and safety of rescue stenting in patients who failed mechanical thrombectomy from our multiethnic stroke patients.

## Materials and Methods

### Participants

Consecutive adult patients (age ≥18 years) with acute stroke patients with an intracranial LVO were retrospectively reviewed from a prospectively collected multiethnic stroke database(15). All patients with acute stroke who underwent thrombectomy and rescue stenting from April 2015 till September 2020 were included in the study. Neurologic inclusion criteria for MT were acute ischemic stroke with an National Institute of Health Stroke scale (NIHSS)(16) score of ≥ 2 and onset time < 24 hours. All patients underwent baseline non-contrast computed tomography (CT) head, CT-angiography (CTA) and CT-perfusion (CTP) before endovascular treatment. Neuroradiological inclusion criteria were Alberta Stroke Program Early CT Score (ASPECTS)(17) ≥ 6 and presence of relatively small core and large penumbra in the extended time window (> 6 hours) or if time of onset is unknown for anterior circulation strokes, lack of bilateral brain stem infarcts for posterior circulation on either CT or diffusion weighted imaging magnetic resonance imaging (DWI MRI), by visual assessment and agreement between the interventionist and the stroke neurologist. Thrombectomy was done if LVO of the following were diagnosed :in the anterior circulation the Internal Carotid artery (ICA) terminus/Middle Cerebral artery (MCA)-M1 and M2) and in the posterior circulation ;the basilar trunk or tip and the Posterior Cerebral artery (PCA)-P1 site on CTA. Digital subtraction angiography and endovascular thrombectomy were performed in all patients.

### Endovascular Procedure

After sedation an 8-Fr balloon-guiding catheter (Cello, Medtronic, USA) was positioned in the ICA for anterior circulation or Neuroon Max 088 (Penumbra Inc, USA) was positioned in the distal vertebral artery for posterior circulation. The intracranial occlusion was crossed with a microcatheter (Rebar 18 or 27, Medtronic, USA). Thrombectomy was attempted with Solitaire AB 4×20 (EV3 Neurovascular, Irvine, California) or Solitaire FR 4×20 (EV3 Neurovascular, Irvine, California) in 87.3% of cases. Infrequently, the Trevo 4×20 (Stryker neurovascular, USA) or Embotrap II 5×30 (Cerenovus, USA) were utilized in the remainder of the cases. The procedure was concluded if there was successful recanalization (Thrombolysis in cerebral infarction (TICI) 2b/3) without residual stenosis. If there was recanalization with residual severe stenosis or immediate re-occlusion post recanalization or failure to recanalize after maximum of 3-5 passes of stent retriever, we considered permanent placement of a stent A Solitaire AB 4 × 20 stent (Medtronic, USA) was used in most of the cases. Only, in 3 patients Leo stent (BALT, France) was used.

Successful recanalization was defined as achieving modified Thrombolysis in cerebral infarction (TICI) scale (TICI)(18) grade 2b or 3 and no reocclusion observed on delayed angiograms during the procedure.

### Postprocedural Antithrombotic Medication and Follow-Up Examinations

After stent detachment, an intra-arterial bolus of Eptifibatide was administered according to the weight of the patient (180 µg/Kg). It was followed by a 24-hours intravenous infusion of Eptifibatide (0.75 μg/Kg/min) for patients with prior tissue plasminogen activator (tPA) treatment and Eptifibatide (2 μg/Kg/min) without prior tPA treatment. After 24 hours a CT was performed to rule-out asymptomatic or symptomatic intra-cerebral hemorrhage (sICH) and CTA was performed to document stent patency, if no bleeding was reported and stent not occluded, dual antiplatelet of were administered (Aspirin 100 mg and Clopidogrel 75 mg). Dual antiplatelet treatment was continued for 3 months.

### Collected Variables

Demographics, Stroke risk factors, stroke severity utilizing the National Institutes of health Stroke Scale (NIHSS) at admission and discharge, the use of tPA, door-to-needle, CT-to-groin puncture and groin puncture to recanalization timings. ASPECTS scores prior to procedure, the collateral scores dichotomized to 0-1 indicating poor collaterals and 2-3 indicating good to excellent collaterals. (19), number of passes. The recanalization score (TICI) categorized as optimal/suboptimal recanalization (TICI =2b-3),poor recanalization (TICI= 1-2a) and occlusions, and the Modified Rankin score (mRS)(20) pre-stroke, at discharge and at 90 days.

#### Primary Outcomes

1) Safety outcome: Incidence of intracerebral hemorrhage. Symptomatic intracranial hemorrhage (sICH) was defined as increase in NIHSS ≥4 with either parenchymal hemorrhage (PH)1()or PH2 on imaging according to The Heidelberg Bleeding Classification (21).
2) Efficacy Outcomes: Radiologic: TICI score and Clinical: mRS at 90 days.

Good clinical outcomes were defined as mRS (0-3) as large vessel occlusions are typically moderate to severe strokes(22).

Secondary Outcomes: Radiologic: Stent Patency at 24 hours; Clinical: NIHSS at discharge. Comparison was made between patients from the Middle East and North Africa (MENA) region and others. The MENA region was defined as per, the United Nations International Children’s Emergency Fund (UNICEF) definition(23),

### Statistical analysis

Standard descriptive statistics were used to present the data. All tests were two-sided at P < 0.05. Analyses were undertaken using SPSS 21.0 (SPSS Inc., Chicago, IL, USA).

### Ethical consideration

This study was a retrospective data analysis approved by institutional review board of the Medical Research Center Protocol # MRC-01-20-071. The authors declare that there is no conflict of interest.

## Results

Out of 220 thrombectomies, 55 (25%) patients underwent rescue stenting with a predominance of male patients (87.3%). The mean age was 51.3±11.8 years. Thirty-two patients (58.3%) were from South Asia, 12 (21.8%) were from the Middle East and North Africa (MENA) region, 9 (16.4%) were from Southeast Asia and 2 (3.6%) were from other parts of the world.

Demographics and risk factors are shown in Table 1.

**Table 1.**
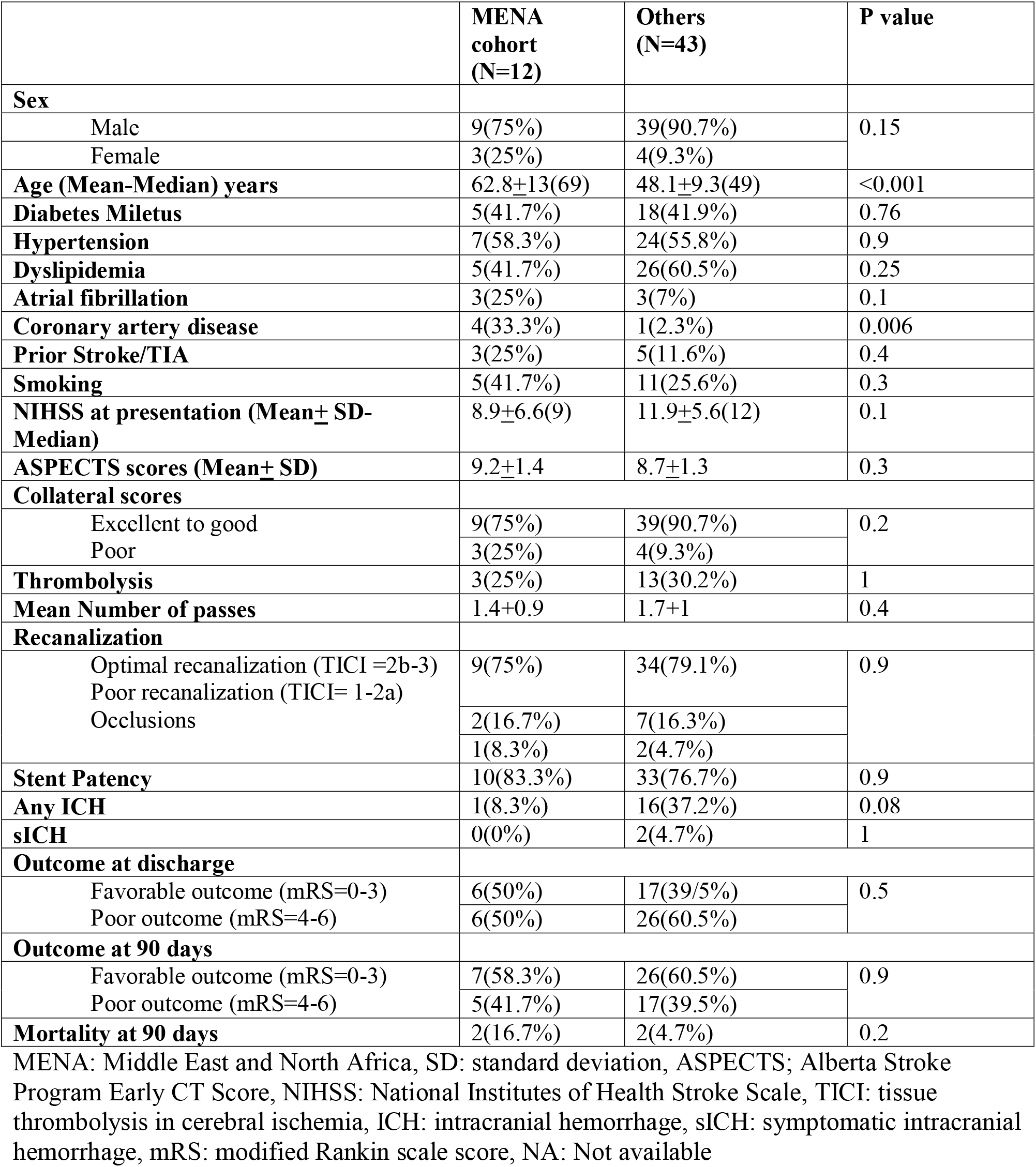
Demographics, clinical characteristics, and outcome differences between patients from the MENA region versus others

Almost all patients had an excellent pre-stroke modified Rankin score (mean 0.04 ± 0.2 median 0). The mean NIHSS score on presentation was 11.3 ± 5.8 (median 11). Table 1 compares between patients from the MENA region vs others. MENA patients were significantly older than others 62.8±13 vs.48.1±9.3 years (p<0.001).

### Pre-Intervention Imaging

Most patients had good to excellent ASPECTS scores (≥7) 51/55 (92.7%). The middle cerebral artery (MCA) M1 segment was the most frequent site of occlusion 65.5%, followed by the supraclinoid portion of the internal carotid artery (ICA) (21.8%) and the MCA M2 segment (7.3%). The majority of patients had good to excellent CT collateral scores (87.3%).

### Thrombolysis & Thrombectomy

Only (29.1%) received both thrombolysis and thrombectomy. Mean door-to-needle time (DTN) was 62.8±45.5 mins (median 41 mins, range 14-171). Mean CT-to-groin time was 96.9 ±33 mins (median 94 mins, range 21-180). The majority of patients (83.6%) received conscious sedation with local anesthesia with the rest receiving general anesthesia. The mean puncture-to-recanalization time was 53.8±23.8 mins (median 49 mins, range 16-125). The mean number of passes was 1.6±1 (median 1, range 1-5). Balloon Angioplasty was performed only in 10 patients (18.2%).

Immediate post rescue stenting residual significant stenosis (50-99%) was present in 25.5% while 7.3% had complete occlusion. There was no intraprocedural complications. Optimal recanalization (TICI =2b-3) was achieved in 43/55(78.2%); whereas 9/55 (16.4%) had poor recanalization (TICI= 1-2a) and 3 (5.5%) had total occlusion post rescue stenting.

### Post procedure management

Post treatment protocol was followed in (91%) of patients. One patient received a loading dose of eptifibatide followed by dual antiplatelets maintenance and one patient received loading with clopidogrel 300mg followed by dual antiplatelets maintenance and 3 patients received no post stenting platelet inhibition due to bleeding on follow up imaging.

### Clinical and radiological Outcomes

The mean NIHSS at discharge 7.1±5.7 (median 7).

Non-invasive angiography (CT or MR) was available for 53/55(96.4%) patients after 24 hours from the procedure. Stents were patent in 44 patients (80%). The two patients who did not have follow up vascular imaging likely both had occlusion as one died from a massive MCA stroke and the other underwent hemicraniectomy due to malignant transformation. Post stenting occlusion was seen in 12/55 patients (21.8%) (10 documented and 2 presumed). Any ICH on 24-hour CT head was seen in 17/55 (30.9%) (hemorrhagic infraction (HI)-1 =5, HI-2=6, Parenchymal hemorrhage (PH)-1=1, PH-2=1, Subarachnoid hemorrhage (SAH)=4). Most ICH patients (15/17; 88.2%) were in the thrombectomy only group without thrombolysis however this was not statistically significant (P=0.06). Only 2/55 (3.6%) patients had symptomatic ICH: one in the thrombectomy only group and one in the thrombectomy + tPA group (p>0.05). Favorable outcome (mRS 0-3) at discharge was seen in 23/55 patients (41.8%), whereas at 90 days, 30/55 (54.5%) had good outcome. Four patients (7.3%) died.

Favorable outcome was seen in 32/43 (74.4%) patients with optimal /suboptimal recanalization, while only in 1/12 (8.3%) with occluded stents at 24 hours (p=0.001). Presence of any ICH on 24-hour imaging was not associated with poor outcome (p=0.08).

Interestingly, apart from significantly older age; mean 62.8±13 years (median 69) vs. 48.1±9.3 (median 49) and coronary artery disease burden 4 (33.3%) vs.1 (2.3%) (p<0.05), patients from the MENA cohort had similar risk factor profiles, stroke severity, recanalization rates, ICH rates and outcomes compared to patients from South and Southeast Asia (Table 1).

## Discussion

To our knowledge, this the first report on rescue stenting in a multiethnic cohort involving patients from the Middle East and North Africa. We report good outcomes and low rates of clinically significant bleeding despite utilization of aggressive platelet inhibition post procedure. Overall, our population seems to be younger than other reported cohorts from China (24), USA(25), Spain(26) and, South Korea(13).

Interestingly, the MENA cohort were older and mirrored the above-mentioned cohorts from USA, Europe and East Asia in age.

Unsurprisingly, our non-MENA cohort (composed mostly of South Asians) mirrored the Indian cohort reported by Sajja et al(27) in age.

Although the presence of hemorrhagic transformation or sICH seemed to be higher in the non-MENA group, this was not statistically significant.

The overall rates of recanalization were comparable to prior reported cohorts (13, 24-27) (Table 2), despite less utilization of angioplasty in our cohort compared to the above-mentioned cohorts. However, the main difference in our cohort is adherence to an aggressive platelet inhibition with glycoprotein IIb/IIIa inhibitors protocol after stenting in the majority of cases (90.9%) while in other cohorts it was left to the treating physician at times or reported in less than 42% of cases. Moreover, we reported lower rates of sICH and mortality than was reported by these cohorts. This could be due to excellent premorbid mRS and lower stroke severity in our cohort (NIHSS median 11 vs 14-17), high ASPECT scores (28), good collateral scores or less endothelial irritation (number of passes)(29).

**Table 2:**
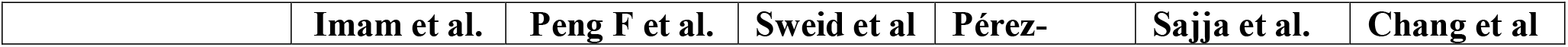
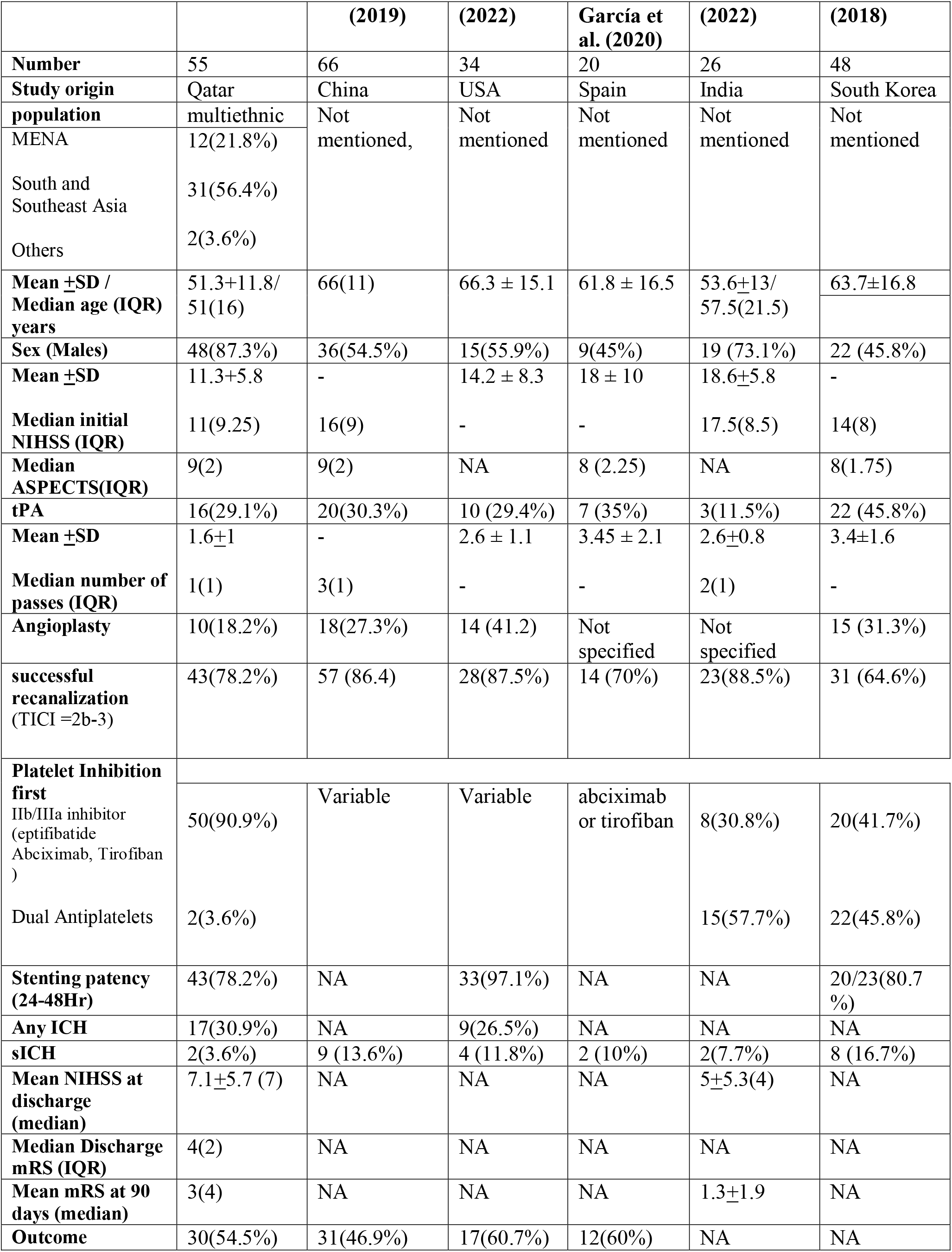
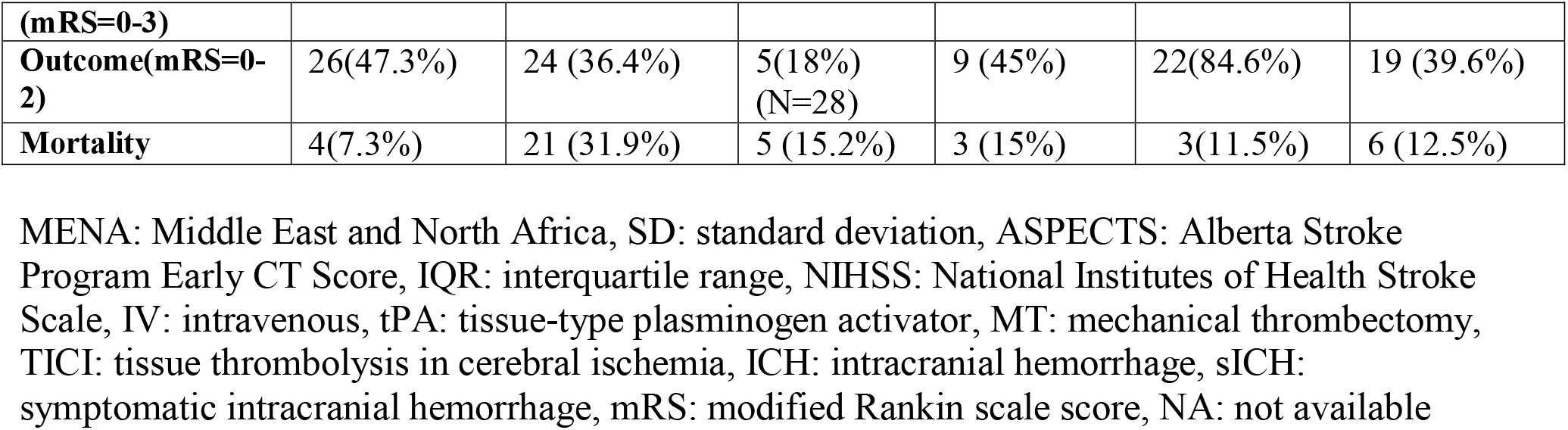
Comparison of different stenting cohorts

Outcomes were similar to reported outcomes in the above cohorts with 54.5% of patients achieving mRS-0-3 and almost half achieving independence, with the only outlier being the Indian study which reported excellent outcomes in 84.6% of cases.

## Conclusion

Rescue stenting showed comparably good outcomes and low rates of clinically significant bleeding in a multiethnic cohort of patients from the MENA and South and Southeast Asia.

## Strength and limitation

The strengths of this study stem from its multiethnic patient diversity, presence of post stenting imaging and antiplatelet protocol.

There are several additional strengths in our study. Firstly, this is the first study on rescue stenting post thrombectomy from a diverse population. Secondly, most of the patients followed a protocol-based management plan. Finally, the presence of some similarities of the MENA cohort with western populations is helpful when compared with the much younger Asian population where ICAD is common. This speaks to possible generalizability of such interventions along with the demonstrated efficacy and safety profile of use of IV eptifibatide in rescue stenting across different cohorts and opens the door for future research.

The main limitation of the study was its retrospective nature. Additionally, there might have been a selection bias of patients and procedural technique as this was dependent on the interventional team.

## Data Availability

All data produced in the present study are available upon reasonable request to the authors

## Notes

### Competing Interest Statement

The authors have declared no competing interest.

### Funding Statement

This study did not receive any funding

### Author Declarations

Ethics committee/IRB of Hamad Medical Corporation gave ethical approval for this work Medical Research Center Protocol MRC-01-20-071.

